# Characterising and differentiating cognitive and motor speed in older adults: a longitudinal birth cohort study

**DOI:** 10.1101/2024.01.04.24300822

**Authors:** Indra Bundil, Sabina Baltruschat, Jiaxiang Zhang

**Affiliations:** School of Psychology, Cardiff University, Cardiff, UK; Department of Computer Science, Swansea University, Swansea, UK

**Keywords:** information processing speed, ageing, cognitive speed, motor speed, letter cancellation, cognitive functions, simple reaction time, choice reaction time

## Abstract

**Objectives:** Information processing speed (IPS) has been proposed to be a key component in healthy ageing and cognitive functioning. Yet, current studies lack a consistent definition and specific influential characteristics. This study aimed at investigating IPS as a multifaceted concept by differentiating cognitive and motor IPS.

**Design, setting and participants:** A retrospective data analysis using data from the Medical Research Council National Survey of Health and Development (MRC NSHD; a population-based cohort of UK adults born in 1946) at childhood (ages 8, 11, and 15) and adulthood (ages 60-64 and 68-70). Using structural equation modelling, we constructed two models of IPS with 2124 and 1776 participants, respectively.

**Outcome measures:** Measures of interest included IPS (i.e., letter cancellation, simple and choice reaction time), intelligence (i.e., childhood intelligence and NART), verbal memory, socio-economic status (SES) and cognitive functions measured by the Addenbrooke’s Cognitive Examination III, as well as a variety of health indexes.

**Results:** We found distinct predictors for cognitive and motor IPS and how they relate to other cognitive functions in old age. In our first model, SES and anti-psychotic medication usage emerged as significant predictors for cognitive IPS, intelligence and smoking as predictors for motor IPS, while both share sex, memory, and anti-epileptic medication usage as common predictors. Notably, all differences between both IPS types ran in the same direction except for sex differences, with women performing better than men in cognitive IPS and vice versa in motor IPS. The second model showed that both IPS measures, as well as intelligence, memory, anti-psychotic and sedative medication usage explain cognitive functions later in life.

**Conclusion:** Taken together, these results shed further light on IPS as a whole by showing there are distinct types and that these measures directly relate to other cognitive functions.

**STRENGTH & LIMITATIONS:**

- A large longitudinal cohort data set with different measurements of information processing speed that are widely used
- Information processing speed is not only related to variables measured at the same time but also to childhood and premorbid intelligence and cognitive functions in later life
- Limitations of the cohort dataset include different response rates between waves, thus some variables were not available for all individuals at certain time points, and IPS scores were derived from a small number of trials
- The study involved self-reported measures which might have increased the proportion of misclassification

## INTRODUCTION

Information processing speed (IPS) is conventionally defined as the speed with which individuals sense, perceive, understand and respond to information (1), and is shown to play a crucial role for cognitive capacity and healthy ageing (2). In the context of the Lothian Birth Cohort (LBC) 1936, previous research found IPS in 70-year olds, measured through Inspection Time (IT), Simple Reaction Time (SRT), and Choice Reaction Time (CRT) as an indicator for intelligence, spatial and verbal abilities (3). Other studies assessing IPS with the same measures, i.e., IT, SRT and CRT, indicated that all three were associated with general cognitive abilities; although only SRT and CRT but not IT related to childhood intelligence (4). Moreover, research on the LBC 1936 and the preceding LBC 1921 confirmed a decline in IPS during ageing (5), suggesting that IPS might serve as a buffer for age-related cognitive decline.

Interestingly, divergent findings emerge when examining the relationship between IPS and education, with studies presenting a varied tapestry of results. While Zhang et al.’s research revealed a significant association using the Digit Symbol Substitution Test (DSST) (6), Ritchie et al. reported a dissociation between education and IPS using LBC 1936 data (7). Regarding sex differences in IPS performance, some studies showed that females are faster, e.g., in tests involving digits and rapid naming tasks (8–10) while others found males to perform better, e.g., in the Finger Tapping Test (FTT) (11) and reaction time tasks (RTTs) (12,13).

As these findings indicate heterogeneous and sometimes even opposite associations; a closer look at the used measures reveals that there are two main approaches to assessing IPS. Firstly, IPS is often measured with classic neuropsychological cognitive batteries such as the Wechsler-Adult-Intelligence-Scale (WAIS) or letter cancellation test (LCT). In these tasks, the crucial outcome variable is performance (e.g., accuracy). In other words, time is constant, i.e., participants have X minutes to work on a task and the accuracy highly differs between individuals as a result.

The second approach often found in studies is assessing response times, that is classical RTTs with varying number of choices and thus difficulty level. Performance is not as important for these tests because, in most cases, there is a ceiling effect (14). Additionally, tasks profoundly differ in the extent to which they capture cognitive and motor processes. However, up to date, there is little research on systematic distinct examinations of these IPS constructs.

In an effort to better understand the differences between them, we attempt to model separate variables to capture cognitive and motor IPS, while acknowledging an overlap in their cognitive and motor components. To deepen this understanding and partially replicate findings from the LBC 1936, our study sets out with two objectives. First, we aim to model individual differences in IPS measures at ages 60-64 (LCT, SRT/CRT) with variables known to be associated with IPS. Second, we aim to study longitudinal association of IPS with cognitive decline; specifically, if both types can predict cognitive functions at ages 68-70, while controlling for the impact of other variables (i.e., health-related, demographic and cognitive). To this end, we employ structural equation modelling (SEM) and use data from the Medical Research Council National Survey of Health and Development (MRC NSHD) cohort, with similar measurements to those of the LBC 1936.

## METHOD

### Cohort data

The MRC NSHD cohort study is a population-based cohort of originally 5362 British-born subjects in March 1946. Data were collected in multiple waves including several ages in childhood (ages 8, 11, and 15) and adulthood (e.g., ages 53, 60-64, and 68-70). Data primarily used in this study are from the waves of 2006-10 (ages 60-64) and 2014-15 (ages 68-70). The time of collection of each individual variable is listed in the Variables section below. To model different types of IPS, we required complete datasets of all IPS variables measured at ages 60-64 (SRT, CRT, LCT), resulting in 2124 cohort members.

The procedure and original protocol have been reported elsewhere (see 15–17) and the data of the MRC NSHD cohort are available to researchers upon request via a standard application procedure. Further details can be found at http://www.nshd.mrc.ac.uk/data.

### Variables

The variables used in the current study are described below. More details on each of these measures can be found in Moulton et al. (17).

#### 1. Information processing speed

##### 1.1 Letter Cancellation Test (LCT)

The task was to detect and cross out the letters “p” and “w” on a sheet of paper amongst many different letters for a duration of one minute (see Figure 1A). Performance was quantified as the number of letters searched. The hit rate (correct letters found) did not need to be considered as a separate measure, as most participants made no errors. This task was assessed at ages 60-64.

**Figure 1.**
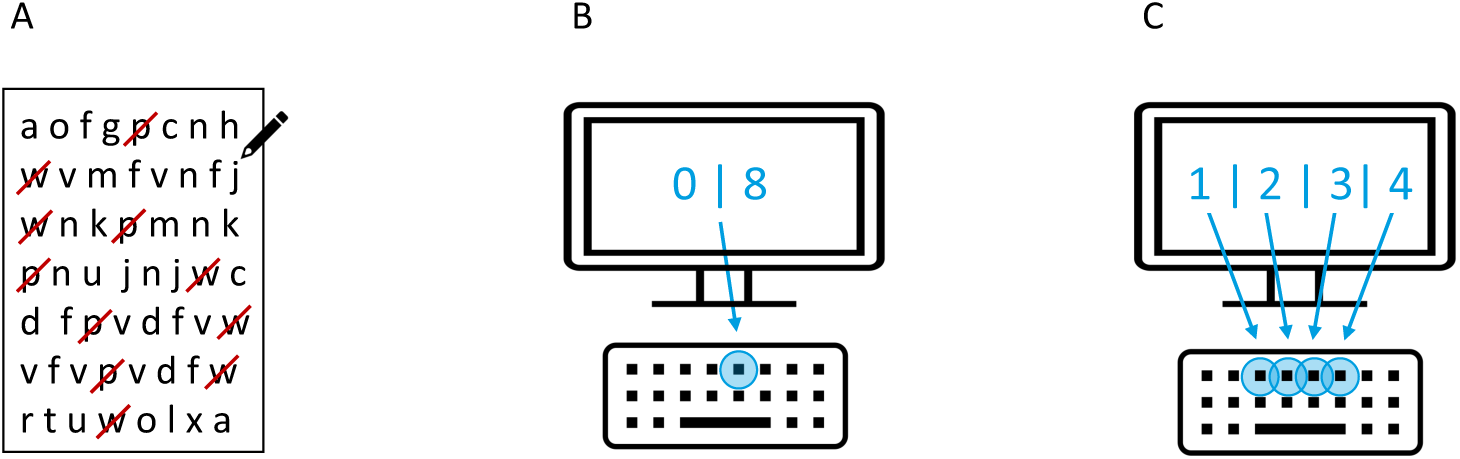
IPS tasks. (A) LCT: Participants detected and crossed out the letters “p” and “w” as quickly as possible, (B) SRT: Participants pressed the same key when the digits “0” or “8” appeared on screen, (C) CRT: Participants pressed the respective keys for digits “1”, “2”, “3”, and “4” appearing on screen.

##### 1.2 Simple Reaction Time (SRT)

A computerised version of the task was used in which participants had to press a specific key as quickly as possible (with only one finger) when the digits “0” or “8” were displayed on the screen (see Figure 1B). After a practice of 8 trials, the main test of 20 trials was used to calculate the mean reaction times (measured in milliseconds). This task was assessed at ages 60-64.

##### 1.3 Choice Reaction Time (CRT)

In the CRT task, one digit from 1 to 4 appeared on screen, and the corresponding key had to be pressed with any finger (see Figure 1C). Again, a practice phase of 8 trials preceded the main test of 40 trials. Mean reaction times were calculated (measured in milliseconds). This task was assessed at ages 60-64.

#### 2. Memory

After being presented a list of 15 words, subjects were asked to write down all words they remembered for one minute. This process was repeated three times. The total score is the sum of all correctly remembered words. This task was assessed at ages 60-64.

#### 3. Intelligence

For childhood intelligence, the standardised summary of all intelligence measures at age 8 was used. If the intelligence scores at age 8 were missing, the measurements from age 11 or 15 were used (in 91 participants). Childhood intelligence measures of all ages differed according to age abilities, but all comprised a cognitive battery entailing picture intelligence (e.g., finding the odd one out or completing a pattern), reading/word comprehension (e.g., completing a sentence with a word out of 5 options), word reading and vocabulary, and arithmetic abilities (for ages 11 and 15 only). For premorbid intelligence, the National Adult Reading Test (NART) was used, measured at age 26 and consisting of the presentation of infrequent words to be read out loud by the participant.

#### 4. Addenbrooke’s Cognitive Examination (ACE-III)

The ACE-III is a battery of neuropsychological tests (18) measuring cognitive functions in five domains: (1) attention and orientation, (2) verbal fluency, (3) memory, (4) language, and (5) visuospatial function. The total score of each domain was used. ACE-III test scores were assessed at ages 68-70.

#### 5. Socio-economic status (SES)

Three variables were used as SES indicators, namely (1) the highest education level reached at 26 years (5 categories: no qualification, vocational only, O-level or equivalent, A-level or equivalent, higher education), (2) the overall social class measured by the type of employment level from ages 26, 36 and 43 (6 categories: professional, intermediate, skilled (non-manual), skilled (manual), partly unskilled, unskilled), and (3) the child social class measured by the employment level of the main income earner (same categories as social class). For this purpose, the values of age 11 or, if not available, age 15 or 4 were used. All social class measures were coded inversely so that a higher class is represented by a lower number.

#### 6. Health indexes

We included several physical health variables, i.e., the most important medications (med.; i.e., anti-psychotic med., sedatives, benzodiazepines, anti-epileptic med., central nervous system (CNS) med., anti-depressants, anti-parkinsonian med. as well as neuromuscular relaxants, BMI, exercise level (3 categories: none, 1-4 times, 5 or more times a month), and smoking status (3 categories: current smoker, ex-smoker, never smoked), all assessed at ages of 60 and 64.

### Pre-processing

To ensure all variables were weighted equally in the model, we pre-processed numerical variables to keep them in a similar range. For each participant, their scores of verbal memory, intelligence, ACE-III, SES and BMI were divided by 10. LCT-scores were divided by 100; SRT and CRT-scores were converted from milliseconds to seconds. We then examined the correlations of all variables (see Table 1) to conceptualise our measurement models and ensure that latent variables were well defined.

**Table 1.** Correlations between all variables used in SEMs.

|  | 1 SRT | 2 | 3 | 4 | 5 | 6 | 7 | 8 | 9 | 10 | 11 | 12 | 13 | 14 | 15 | 16 |
| --- | --- | --- | --- | --- | --- | --- | --- | --- | --- | --- | --- | --- | --- | --- | --- | --- |
| **2 CRT** | .50** |  |  |  |  |  |  |  |  |  |  |  |  |  |  |  |
| **3 LCT** | -.14** | -.25** |  |  |  |  |  |  |  |  |  |  |  |  |  |  |
| **4 Education** | -.17** | -.26** | .16** |  |  |  |  |  |  |  |  |  |  |  |  |  |
| **5 SES** | .16** | .24** | -.14** | -.50** |  |  |  |  |  |  |  |  |  |  |  |  |
| **6 Child SES** | .15** | .18** | -.12** | -.38** | .29** |  |  |  |  |  |  |  |  |  |  |  |
| **7 NART** | .21** | .31** | -.16** | -.54** | .42** | .35** |  |  |  |  |  |  |  |  |  |  |
| **8 Child intelligence** | -.18** | -.29** | .16** | .52** | -.41** | -.36** | -.66** |  |  |  |  |  |  |  |  |  |
| **9 Memory** | -.20** | -.31** | .19** | .39** | -.33** | -.27** | -.48** | .46** |  |  |  |  |  |  |  |  |
| **10 ACE fluency** | -.25** | -.29** | .18** | .27** | -.21** | -.21** | -.38** | .37** | .37** |  |  |  |  |  |  |  |
| **11 ACE language** | -.18** | -.19** | .10* | .28** | -.25** | -.23** | -.44** | .38** | .29** | .30** |  |  |  |  |  |  |
| **12 ACE attention** | -.08* | -.09** | .06* | .17** | -.15** | -.08* | -.17** | .19** | .18** | .20** | .13** |  |  |  |  |  |
| **13 ACE memory** | -.14** | -.22** | .16** | .30** | -.26** | -.23** | -.41** | .36** | .47** | .30** | .35** | .16** |  |  |  |  |
| **14 ACE visuospatial** | -.17** | -.24** | .11** | .30** | -.23** | -.21** | -.30** | .30** | .27** | .26** | .25** | .20** | .30** |  |  |  |
| **15 Exercise** | -.08** | -.11** | .08** | .18** | -.18** | -.14** | -.17** | .17** | .19** | .11** | .09** | .06* | .12** | .09** |  |  |
| **16 Smoking** | -.02 | -.12** | .09** | .15** | -.13** | -.08* | -.11** | .07* | .14** | .05 | .06* | -.02 | .13** | .04 | .12** |  |
| **17 BMI** | .06* | .07* | -.07* | -.12 | .10 | .16 | .11 | -.08 | -.12 | -.07 | -.05 | -.02 | -.07 | -.09 | -.11 | <.01 |

*Note.* *P*-values are highlighted as \* < .05; \*\* < .001;. SRT: Simple Reaction Time, CRT: Choice Reaction Time; LCT: Letter Cancellation Test; SES: Socio-economic Status; NART: National Adult Reading Test; ACE: Addenbrooke's Cognitive Examination

Unsurprisingly, SRT and CRT correlated strongly while LCT only correlated moderately with both RTTs, indicating LCT to represent a different type of IPS than SRT and CRT. We thus used LCT performance as a measure of cognitive IPS, and the SRT and CRT tasks as measures of motor IPS.

Notably, both RTTs include cognitive components, just as LCT includes motor components. Thus, we cannot claim that LCT exclusively measures cognitive and SRT and CRT exclusively measure motor IPS; new task designs would have to be developed for this. However, using SEM, we were able to form latent constructs based on the shared variance between the measured variables. While cognitive processes are required to different degrees for SRT and CRT, their high correlation should reflect their similarity of motor responses.

Since our latent motor IPS variable is constructed from both RTTs, it should thus reflect motor rather than cognitive processes. Cognitive IPS, solely modelled by LCT, still includes some motor responses, but primarily represents cognitive processes that are not represented by the latent motor IPS variable.

### Statistical analyses

We used SEM in lavaan (R package) (19) to test how cognitive and demographic measures relate to cognitive and motor IPS, introduced as latent variables. Mardia and Henze-Zirkler tests showed that the data did not form a multivariate normal distribution; thus, we used maximum likelihood with robust standard errors (MLR) to estimate the latent constructs and the path coefficients of our two models.

Model 1 aimed to explore the predictors of cognitive and motor IPS measured at ages 60-64. Model 2 explored the indicative value of these two IPS measures on cognitive functions at ages 68-70 (i.e., ca. seven years after the IPS measures) measured with the ACE-III battery. For the second model, we only included participants with complete ACE-III measurements, reducing the sample to 1776 cases. Both measurement models with all latent constructs are shown in Figure 2.

**Figure 2.**
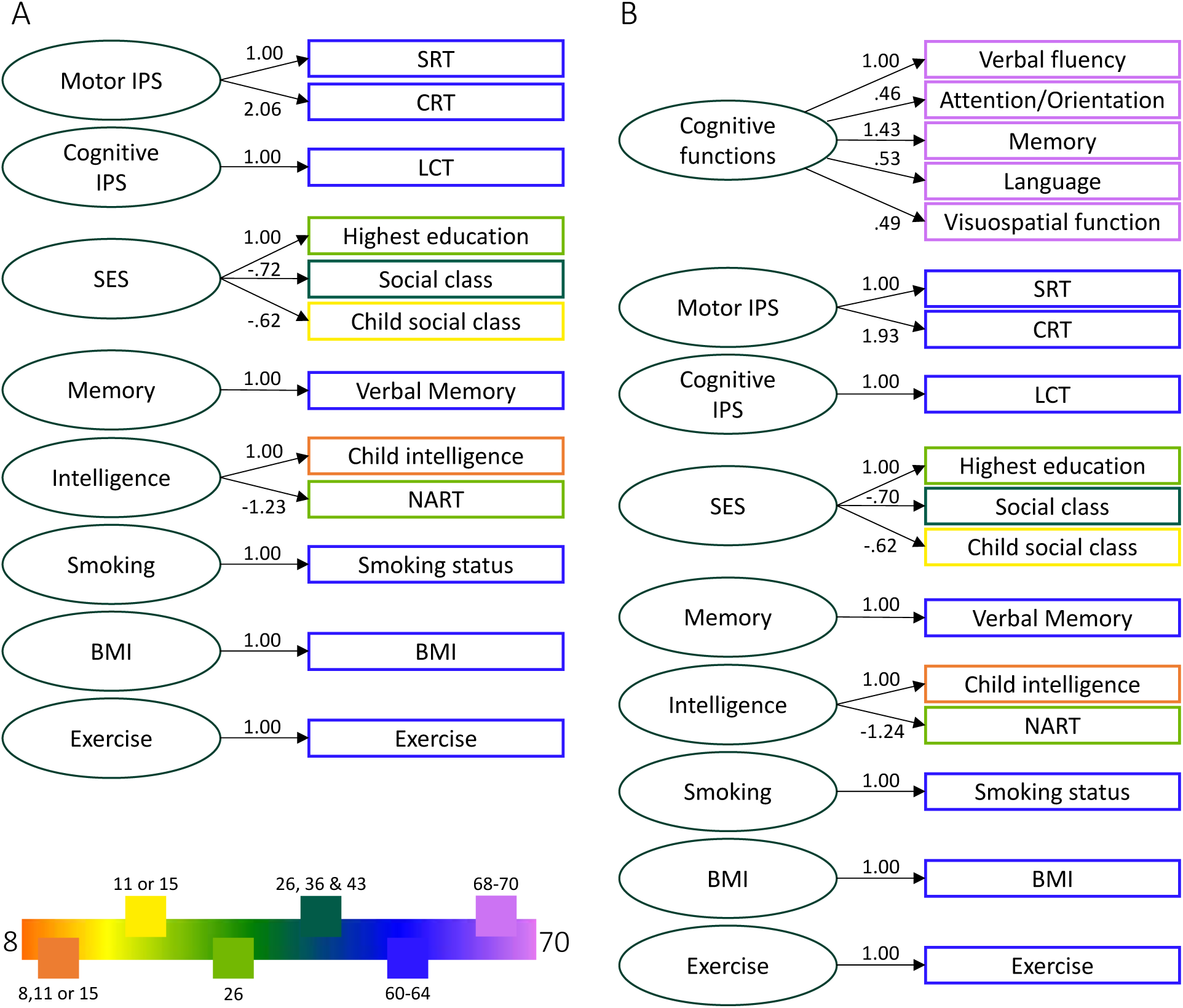
Measurement models for (A) Model 1 and (B) Model 2 with factor loadings. The legend refers to the age of assessment of the respective coloured variables. BMI: Body Mass Index; CRT: Choice Reaction Time; LCT: Letter Cancellation Test; NART: National Adult Reading Test; SES: Socio-economic Status; SRT: Simple Reaction Time.

Cognitive IPS, memory, and confounders (BMI, exercise level, smoking) were included as latent variables with just one predictor as recommended by Hayduk & Littvay (20). Their error terms were set to 0 with lavaan’s single predictors option. Binary variables (sex and medication usage) were introduced in the regression of the path analyses (not in the measurement model). Correlations between all latent variables can be found in the Supplementary Material (Supplementary Tables 2 & 3).

The two structural models were constructed as follows:

Model 1:

*Cognitive/Motor IPS = Socio-economic status + sex + intelligence + memory + smoking + BMI + exercise + CNS med. + benzodiazepines + anti-psychotic med. + anti-depressants + anti-epileptic med. + anti-parkinsonian med. + neuromuscular relaxants + sedatives*

Model 2:

*Cognitive functions = motor IPS + cognitive IPS + socio-economic status + sex + intelligence + memory + smoking + BMI + exercise + CNS med. + benzodiazepines + anti-psychotic med. + anti-depressants + anti-epileptic med. + anti-parkinsonian med. + neuromuscular relaxants + sedatives*

Missing variables were estimated with the maximum likelihood function, as recommended (21,22) (See Supplementary Table 1 for number of missing variables). Model fits were determined by two measures: robust Comparative Fit Index (CFI), which is a score comparing the model to a baseline model and is regarded as acceptable if it is > .9 and good if it is > 0.95 (23); Root Mean Square Error of Approximation (RMSEA) and Standardized Root Mean Square Residual (SRMR), which are absolute measures of fit and are acceptable if they are < .08 and good if they are < .06 (24). The χ2 is also reported for information, but cannot be used as a fit measure as it is highly sensitive to sample size (25) and assumes multivariate normality, rejecting models that do not meet this criterion (26).

As commonly used in SEM research, we use the term “predict” in this study to interpret the results. While this denotes that a variable is a strong marker for another, it is important to clarify that it signifies an observed statistical relationship and should not be construed as implying causality.

## RESULTS

When interpreting the results, it is worth noting that SRT and CRT measure reaction times, i.e., lower times refer to greater IPS, while LCT measures the number of crossed-out words, i.e., a greater number means greater IPS. Thus, contrary directions of cognitive and motor IPS in the models and correlations are showing a relation in the same direction with the variable.

The factor loadings of all variables are shown in Figure 2. Both measurement models have good fit indexes (Model 1: χ2(31) = 62.611, p < .001; RMSEA = .022; SRMR = .014; CFI = .994; Model 2: χ2 (88) = 226.092, p < .001; RMSEA = .031; SRMR = .023; CFI = .975).

Model 1 shows good absolute fit indexes and acceptable comparative fit (χ2(173) = 635.861, p < .001; RMSEA = .038; SRMR = .031; CFI = .929). Sex, memory, and anti-epileptic med. usage emerged as common predictors for both IPS types. Further, motor IPS was predicted by intelligence and smoking, while cognitive IPS was predicted by SES and anti-psychotic med. usage (see Figure 3A). Of note, sex was inversely related to both concepts, with women showing greater cognitive and men greater motor IPS. Moreover, these results need to be interpreted while considering the correlation matrix (see Table 1). For instance, in Model 1, SES but not intelligence emerged as a significant predictor for cognitive IPS (as shown in Figure 3A). However, LCT and both intelligence measures are significantly related with *r* = |.16|. Due to the nature of SEM, intelligence is not significant in Model 1 as it does not explain any further variance of cognitive IPS beyond that what SES already explains.

**Figure 3.**
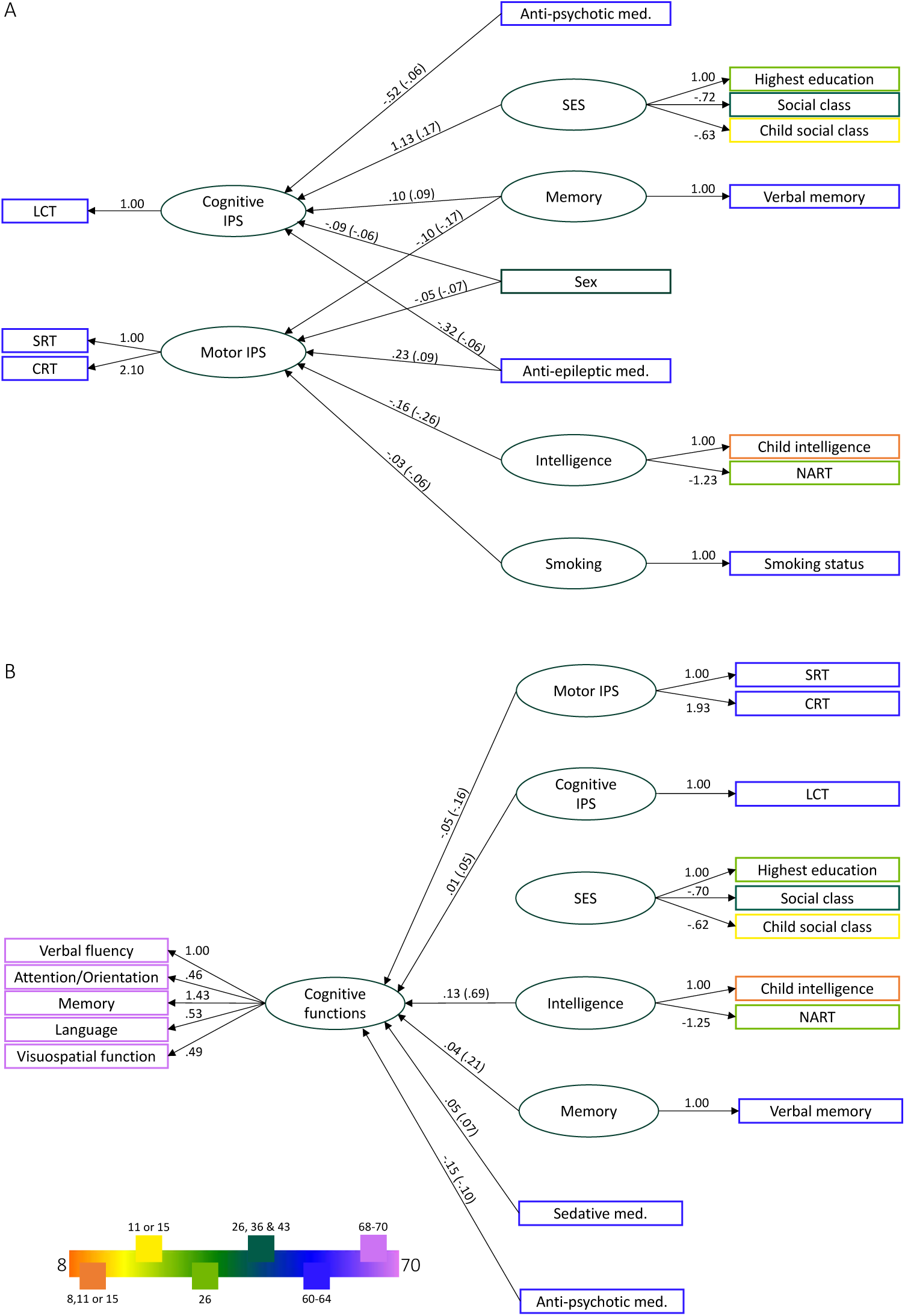
Factor loadings, path coefficients and standardised parameters of both SEMs. A shows all latent variables with their loadings and all significant predictors of Cognitive and Motor IPS (Model 1); B shows all latent variables with their loadings and all significant predictors of cognitive functions measures at ages 68-70 (Model 2). Standardised parameters are noted in parenthesis. The legend refers to the age of assessment of the respective coloured variables. BMI: Body Mass Index; CRT: Choice Reaction Time; LCT: Letter Cancellation Test; Med: Medication; NART: National Adult Reading Test; SES: Socio-economic Status; SRT: Simple Reaction Time.

Model 2 also shows a good absolute and acceptable comparative fit (χ2(248) = 739.414, p < .001; RMSEA = .034; SRMR = .032; CFI = .916). Both IPS measures, as well as intelligence, memory, anti-psychotic and sedative med. usage significantly predicted cognitive functions measured seven years later (see Figure 3B).

To ensure that the imputation of missing data is not causing biased results, we repeated the analyses of the models only with complete data sets; i.e., 1600 cases for Model 1 (75% of the participants of the previous model) and 1156 cases for Model 2 (65% of the participants of the previous model). With the exception of SES no longer being significant for cognitive IPS and only anti-parkinsonian med. emerging as a predictor among the medications, Model 1’s results are robust. For Model 2, cognitive IPS is a weaker and no longer significant predictor. These additional analyses indicate that our main results are robust to the imputation of missing data. Details of these analyses can be found in the Supplementary Material (also see Supplementary Figure 1).

## DISCUSSION

The present study reported that, among older adults, cognitive IPS (measured by the LCT) and motor IPS (measured by RTTs) are two constructs that are predicted by different cognitive and demographic variables. Both, cognitive and motor IPS, overlap in having memory, sex, and anti-epileptic med. as common predictors. Motor IPS is further predicted by intelligence and smoking, and cognitive IPS is further predicted by SES. Using a different SEM implementation, we observed that cognitive and motor IPS predict cognitive functions (measured by the ACE-III) seven years later.

It is important to note that, although some variables have pairwise correlations with cognitive or motor IPS, they are not significant predictors in the SEMs (e.g., exercise, BMI, CNS med., benzodiazepines, anti-depressants, anti-parkinsonian med., and neuromuscular relaxants). This is because such variables do not explain any further variance of the endogenous variables that is not already explained by other predictors or latent variables. Below, we focus our discussions on the significant variables from the SEMs.

### Cognitive and motor IPS measures differ at the theoretical and neural level

At the theoretical level, cognitive IPS can be defined as the rate at which perceptual and automatic cognitive tasks are executed. Cognitive IPS is commonly tested in tasks requiring a sustained level of attention under time constraints (27,28). The LCT from the MRC NSHD requires participants to detect and cross out specific letters, entailing the memorisation of target letters and the discrimination of such against distractor letters, while requiring verbal abilities along the way. This puts a high focus on cognitive abilities, linguistic or visuospatial abilities and lower-order abstraction skills (29). Motor IPS as assessed by RTTs, on the other hand, focuses on the efficiency of speeded motor responses (30). Contrary to the LCT, the behavioural responses in RTTs only involve simple motor processes (i.e., pressing specific buttons with specific fingers), from which reaction times statistics can be measured.

There are cognitive components in both SRT and CRT, as well as motor components in LCT. Both RTTs involve visual scanning, discriminating the displayed stimuli and deciding on the appropriate motor response. Moreover, in CRT, participants must decide not only when to make a response but also which response to make (i.e., make a choice among response options). Therefore, CRT involves more extensive cognitive processes than SRT, which is reflected in the higher correlation of LCT with CRT than with SRT (see Table 1). Conversely, LCT captures visuomotor processes as letters must be scanned across the paper (as opposed to constant eye fixation on the screen in computerised RTTs) and hand-eye coordination as letters are crossed. Nevertheless, it is important to point out that the motor IPS variable in the current study is formed by the common variance of CRT and SRT, which may be primarily related to the motor components of the two tasks, which are different from those in the LCT.

At the neural level, motor and cognitive IPS tasks differ in the extent to which they require secondary association areas (involved in perceptual analysis and synthesis, object recognition, (non)linguistic analysis, and preparation of motor actions) (29). Cognitive IPS, in particular high LCT performance, is related to increased activity in the middle frontal gyrus, and reduced activity in the cerebellum (31). Better performance in motor IPS, as measured by SRTs, was previously found to be mostly associated with activation in the occipital lobes, sensorimotor cortices, and supplemental motor cortices (32) along the sensorimotor pathway (33).

Altogether, our findings confirm the theoretical distinction between cognitive and motor IPS. While common predictors reflect the shared variance of motor and cognitive IPS (such as sensation and perception), distinct predictors underlie the theoretical and neural differences stated above.

### Demographic factors: sex and SES

Model 1 showed women to perform better than men in cognitive IPS and vice versa in motor IPS tasks which might explain previous heterogeneous research as stated earlier. Further studies consistently showed that females outperform males in tests involving digits and rapid naming tasks, thus in cognitive IPS tasks. This may be due to sex differences in phonological coding involved in speech-based processes (i.e., reading and writing abilities; for a systematic review see 34). Specifically, sex differences in performance were detected in tasks with letters(-strings) (35,36), but not in tasks involving geometrical figures (9). Consistent with our results, O’Shea et al. found women to perform higher in the LCT (37).

Considering motor IPS, research using the FTT (11) (for a review see 38) and RTTs consistently showed that men are faster than women (12,13), consistent with the results in the current study. According to Reimers & Maylor, variability in RTTs possibly reflects sex differences in the trial-to-trial speed-accuracy trade-off, as they found that women were slower and more accurate than men at the beginning but became faster later in test sessions (39). However, as the SRT and CRT tasks in the NSHD dataset consisted of a small number of trials with no trial-to-trial data, we were not able to confirm this proposition in the current study.

In a study of the Aberdeen Birth Cohort (ABC) of 1936, SES, composed of parental and own occupational status, predicted late life IPS (40). However, they also found that education and childhood ability were better predictors. Similarly, we found that SES including parental and own occupational status, as well as education level, could predict cognitive IPS and that childhood intelligence is a better predictor for motor IPS. The high covariance between our intelligence and SES latent variables highlights the close relationship and supports the shared variance described by Staff et al. (40).

The difference in the relationship of our IPS measures and SES may, again, be a result of the different tasks. The DSST used in the ABC is similar to the LCT, but does not involve linguistic processing. As in the ABC study, other research found a correlation between the DSST and SES (6,41). Contrastingly, in the English Longitudinal Study of Aging (ELSA), SES was not correlated with cognitive IPS, measured with the LCT at age 63 (42), but they found that a greater decline in cognitive IPS over time was associated with lower SES. The differences in how SES was modelled may have led to the distinct results. Nevertheless, we can conclude that this study is in line with our findings, as the relation of SES and decline in cognitive IPS was negative, pointing to a negative impact of lower SES on cognitive IPS.

### Cognitive factors: intelligence and memory

Faster IPS has been related to higher intelligence, especially better g-factor or general intelligence scores (2,43). In a previous study, FTT performance was positively correlated to intelligence as measured by the Raven’s progressive matrices, WAIS-III verbal IQ, and full-scale IQ (11). This is in line with our results as we found that motor IPS was predicted by the latent variable intelligence in Model 1. Further, studies on the LBC 1936 confirmed that SRT and CRT are moderately related to childhood and premorbid intelligence, measured with the NART (3,5), just as in the present study. Other research on this cohort also showed, that RTTs were related to general intelligence and that this correlation becomes more pronounced as the number of response options increases (10,44). This, again, is in line with our results, as the correlations of child and premorbid intelligence with CRT are greater than with SRT (44).

In this way, we replicated previous findings of the association between motor IPS and intelligence. However, our cognitive IPS variable measured with the LCT shows a lower correlation with intelligence variables and is not predicted by the intelligence latent variable in our model. Thus, our results point to different characteristics between motor and cognitive IPS. In fact, less work was done on the relationship between cognitive IPS and intelligence and results are heterogeneous with some studies not finding a direct correlation to intelligence at all (45).

In previous literature, IPS was mostly related to working memory, which is different to short-term memory measured in the MRC NSHD cohort (46–49). The research measuring short-term memory found moderate correlations between memory tasks and some of the IPS measures (49–51). Colom et al. (51) and Dang et al. (49), for instance, found that a task, where participants had to cross out numbers, similar to the LCT, showed moderate correlations with visuo-spatial memory and low correlations with verbal memory. This may be due to the domain of the tasks, with a greater relation of visuo-spatial to numeric than to verbal abilities. Conway et al. found that IPS, measured with a simple pattern and letter comparison task, was moderately correlated to verbal memory, but only in an articulatory suppression condition (50). These two studies used several IPS, short-term memory, and working memory tasks confirming that working memory shows a closer relation to IPS than short-term memory. However, with great differences in the instruments used and heterogeneity in the results of each test, it is difficult to draw conclusions on how strong the relation of the different types of IPS and memory are.

### Health factors: smoking and medication

Although the immediate effect of smoking, i.e., high levels of nicotine, has been related to better cognitive functions (52), long-term smokers have shown worse performance in cognitive tasks in later life (53–55). For instance, a faster decline in memory in later life was found in both the LBC and ABC cohorts (54,55). In terms of IPS measures, cognitive IPS was reported to be decreased in smokers of the LBC (54) and the DSST also showed lower performance in smokers (55). However, in general, effect sizes have been low, just as in our study. We also found smoking as a predictor for motor IPS and not cognitive IPS as in the LBC. This may be due to age differences at the time of data collection and in the design of the statistical models, as the raw correlations of all IPS measures with smoking status are low.

We found that anti-epileptic, anti-psychotic, and sedative med. were significant predictors of IPS. Previous research found that epilepsy is linked to white matter loss and slower reaction times (56,57), and some anti-epileptic drugs have been reported to affect cognition and IPS (58). In psychotic disorders, it is known that most patients already have cognitive deficits before the first psychotic episode (59). More specifically, schizophrenia is associated with IPS impairments using the DSST, as shown by a meta-analysis (60).

Sedatives have also been associated with cognitive impairment. Depending on the drug, different cognitive processes are affected, e.g. memory (61), as well as attention and psychomotor functions (62). In our study, however, we found a positive effect of sedatives on cognitive functions measured in later life. This could be due to the variety of reasons for the use of these drugs, as they are also taken for sleep disorders and are not necessarily associated with a disease that involves cognitive impairment.

### IPS as a predictor of cognitive functions in later life

Model 2 explored understanding the temporal dynamics of motor and cognitive IPS on cognitive functions 7 years later. Our results showed that later-life cognitive functions, measured by the ACE III at ages 68-70, are predicted by both motor and cognitive IPS. There is a large body of research on the cross-sectional association between IPS and cognitive functions (63–65), but only a few investigated longitudinal effects. Finkel et al. applied growth curve modelling of IPS and cognitive functions, showing that faster IPS relates to slower deterioration in spatial and memory performance (66). Taken together, our results are in line with the general notion that IPS is strongly associated with cognitive functioning.

These findings have implications for understanding the stability of cognitive abilities over time. The factors captured by motor and cognitive IPS seem to be important contributors to cognitive abilities that remain consistent over a 7-year period. This could suggest that interventions or strategies targeting these factors (e.g., in the context of cognitive training) might have the potential to lead to long-term improvements in cognitive abilities.

### Implications and future directions

IPS measures, such as the LCT and RTTs, as well as the Colour Trails Test and DSST are frequently used in clinical settings for the diagnosis of patients. Our results show that the performance in such tests cannot be interpreted as a single IPS construct, because cognitive and motor IPS relate to different cognitive and demographic factors. We used data from the MRC NSHD cohort, allowing us to base our findings on a large longitudinal cohort. As all waves include a medical and cognitive assessment, we were able to consider confounder variables, such as medication usage and smoking status. This allowed us to design SEM analyses with important predictors of IPS, as well as other variables that could have influenced the measurements. However, two issues require further consideration. First, we were constrained by the available variables in the NSHD dataset, in which other forms of IPS measures (e.g., DSST) were not assessed. Second, motor IPS was estimated from a limited number of trials as a part of a complicated protocol. As a result, we could not apply cognitive modelling for motor IPS (67) beyond simple RT statistics. Future studies should consolidate our findings in other cohort datasets and IPS tasks.

## CONCLUSION

In conclusion, our results supported a multifaceted concept of IPS, as motor and cognitive IPS are predicted by different variables in older adults. Motor IPS is a predictor of later life cognitive functions, while cognitive IPS is a weaker predictor. These results support the use of IPS as one of the fundamental markers of intelligence and cognition, although differences in task design must be taken into account.

## Acknowledgement

We thank the study participants for their continuing participation in the MRC NSHD and also members of the scientific and data collection teams who have been involved in the data collection.

## Data availability statement

The data of the MRC NSHD cohort are available to bona fide researchers upon request via a standard application procedure. Further details can be found at http://www.nshd.mrc.ac.uk/data (doi: 10.5522/NSHD/Q101; doi: 10.5522/NSHD/Q102; doi: 10.5522/NSHD/Q103).

## Patient consent for publication

Not applicable.

## Funding

This project has received funding from the European Research Council (ERC) under the European Union’s Horizon 2020 research and innovation programme (grant agreement No. [716321 - FREEMIND]). IB is supported by a PhD studentship from Cardiff University School of Psychology.

## Competing interests

None declared.

## Author contributions

JZ and SB conceptualised the study and designed the analyses. SB and IB analysed the data. SB and IB drafted the manuscript. JZ revised the manuscript. All authors reviewed, edited, and approved the final manuscript.

## Patient and public involvement

Patients and/or the public were not involved in the design, or conduct, or reporting, or dissemination plans of this research.

## Notes

### Competing Interest Statement

The authors have declared no competing interest.

### Funding Statement

This project has received funding from the European Research Council (ERC) under the European Unions Horizon 2020 research and innovation programme (grant agreement No. [716321 - FREEMIND]). IB is supported by a PhD studentship from Cardiff University School of Psychology.

### Author Declarations

This study used pre-collected data from the he Medical Research Council (MRC) National Survey of Health of Development (NSHD) cohort. The data access request was approved by the NSHD data sharing committee (project number: JZ1121) upon request via a standard application procedure. Further details can be found at http://www.nshd.mrc.ac.uk/data (doi: 10.5522/NSHD/Q101; doi: 10.5522/NSHD/Q102; doi: 10.5522/NSHD/Q103).

