## Supplementary Material for "Characterising and differentiating cognitive and motor speed in older adults: a longitudinal birth cohort study"

**Supplementary Table 1.** Number of missing data for individual variables, separately for Model 1 and Model 2

| | **Model 1** (*N*=2124) | **Model 2** (*N*=1776) |
|---|---|---|
| **CNS med.** | 1 | - |
| **Anti-psychotic med.** | 1 | - |
| **Anti-depressants** | 1 | - |
| **Anti-epileptic med.** | 1 | - |
| **Anti-parkinsonian med.** | 1 | - |
| **Sedatives** | 1 | - |
| **Highest education level** | 18 | 12 |
| **Overall social class** | 11 | 6 |
| **Child social class** | 109 | 93 |
| **Exercise** | 63 | 45 |
| **Smoking** | 191 | 127 |
| **BMI** | 3 | 2 |
| **Child IQ** | 147 | 123 |
| **NART** | 158 | 108 |
| **Memory** | 38 | 19 |

*Note.* BMI: Body Mass Index; Med: Medication; NART: National Adult Reading Test

**Supplementary Table 2.** Correlations of latent variables for Model 1

| | Motor IPS | Cognitive IPS | SES | Intelligence | Memory | Smoking | BMI |
|---|---|---|---|---|---|---|---|
| **Cognitive IPS** | -.3** | | | | | | |
| **SES** | -.476** | .249** | | | | | |
| **Intelligence** | -.502** | .218** | .932** | | | | |
| **Memory** | -.402** | .199** | .582** | .646** | | | |
| **Smoking** | -.142** | .093** | .223** | .129** | .148** | | |
| **BMI** | .084** | -.067* | -.199** | -.132** | -.122** | .003 | |
| **Exercise** | -.142** | .081* | .289** | .235** | .191** | .132** | -.109** |

*Note.* * < .005; ** < .001

**Supplementary Table 3.** Correlations of latent variables for Model 2

| | **Motor IPS** | **Cognitive IPS** | **SES** | **Intelligence** | **Memory** | **Smoking** | **BMI** | **Exercise** |
|---|---|---|---|---|---|---|---|---|
| **Cognitive IPS** | -.324*** | | | | | | | |
| **SES** | -.528*** | .228*** | | | | | | |
| **Intelligence** | -.586*** | .212*** | .933*** | | | | | |
| **Memory** | -.441*** | .177*** | .574*** | .632*** | | | | |
| **Smoking** | -.166*** | .082*** | .22*** | .124*** | .133*** | | | |
| **BMI** | .101*** | -.056* | -.196*** | -.116*** | -.114*** | -.026 | | |
| **Exercise** | -.121*** | .061* | .267*** | .209*** | .168*** | .129*** | -.095*** | |
| **Cognitive functions** | -.674*** | .292*** | .834*** | .924*** | .734*** | .146*** | -.129*** | .215*** |

*Note.* * < .05; ** < .005; *** < .001

### Results of analysis without imputing missing data

The analyses of the models only with the cases with complete data show very similar results with acceptable comparative and good absolute fit measure: Model 1 ($\chi^2$ = 442.473, *df* = 131, $p < .001$; RMSEA = .038; SRMR = .033; CFI = .927) and Model 2 ($\chi^2$ = 645.878, *df* = 248, $p < .001$; RMSEA = .038; SRMR = .036; CFI = .9). Supplementary Figure 1 shows the models and the factor loadings and path coefficient and $R^2$.

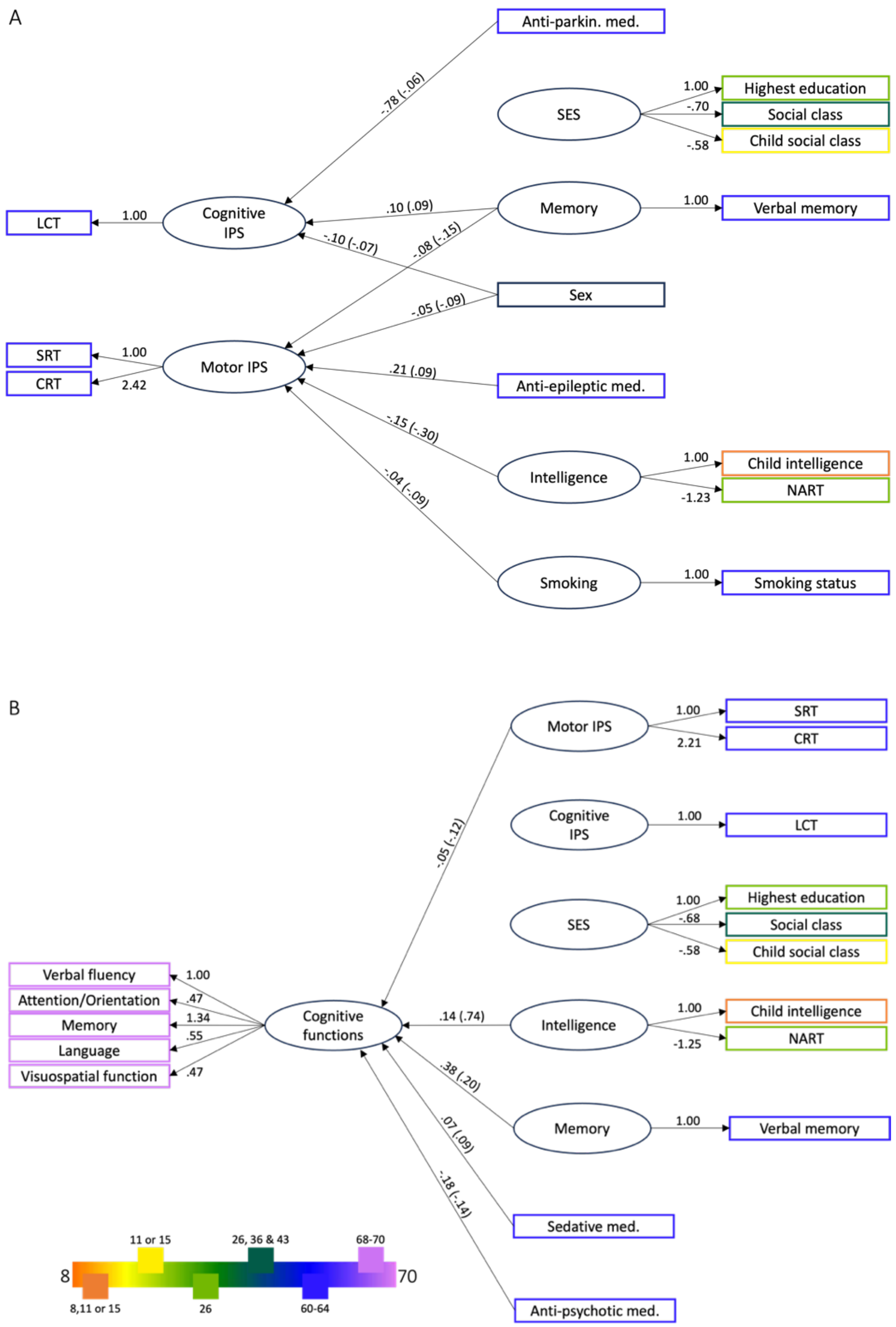


**Supplementary Figure 1. Factor loadings, path coefficients and standardised parameters of both SEMs without imputing missing data.** A shows all latent variables with their loadings and all significant predictors of cognitive and motor IPS (Model 1); B shows all latent variables with their loadings and all significant predictors of cognitive functions measures at ages 68-70 (Model 2). Standardised parameters are noted in parenthesis. The legend refers to the age of assessment of the respective coloured variables. BMI: Body Mass Index; CRT: Choice Reaction Time; LCT: Letter Cancellation Test; Med: Medication; NART: National Adult Reading Test; Anti-parkin.: Anti-parkinsonian; SES: Socio-economic status; SRT: Simple Reaction Time.
